# Anteroposterior balance reactions of children with and without cerebral palsy: a cross-sectional, observational study of children age 5-12 years

**DOI:** 10.1101/19006585

**Authors:** Jeremy R. Crenshaw, Drew A. Petersen, Benjamin C. Conner, James B. Tracy, Jamie Pigman, Henry G. Wright, Freeman Miller, Curtis L. Johnson, Christopher M. Modlesky

## Abstract

**Aim:** To compare anterior and posterior standing balance reactions, as measured by single-stepping thresholds, of children with and without cerebral palsy (CP).

**Method:** Seventeen ambulatory children with CP and 28 typically developing children, all 5-12 years of age, were recruited for this cross-sectional, observational study. Balance reaction skill was quantified as anterior and posterior single-stepping thresholds, or the treadmill-induced perturbations that consistently elicited a step in that direction. In order to understand underlying mechanisms of between-group differences in stepping thresholds, dynamic stability was quantified using the minimum margin of stability. Ankle muscle activation latency, magnitude, and co-contraction were assessed with surface electromyography.

**Results:** We observed large between-group differences in anterior, but not posterior, thresholds. Between-group differences were most evident in older children, with typically developing children having larger anterior thresholds than those with CP. In response to near-threshold anterior perturbations, older typically developing children recovered from more instability than their CP peers. Older children had no between-group differences in ankle muscle activity.

**Interpretation:** The effects of CP on balance reactions are age- and direction-specific. In response to an anterior perturbation, older typically developing children recovered from more instability than their peers with CP.

**What this paper adds:**

- Children with CP have age- and direction-specific balance-reaction impairments.
- Impairment was most evident in the anterior reactions of older children (≈11 years).
- Typically developing children recovered from more anterior instability than those with CP.

**Graphical Abstract:** 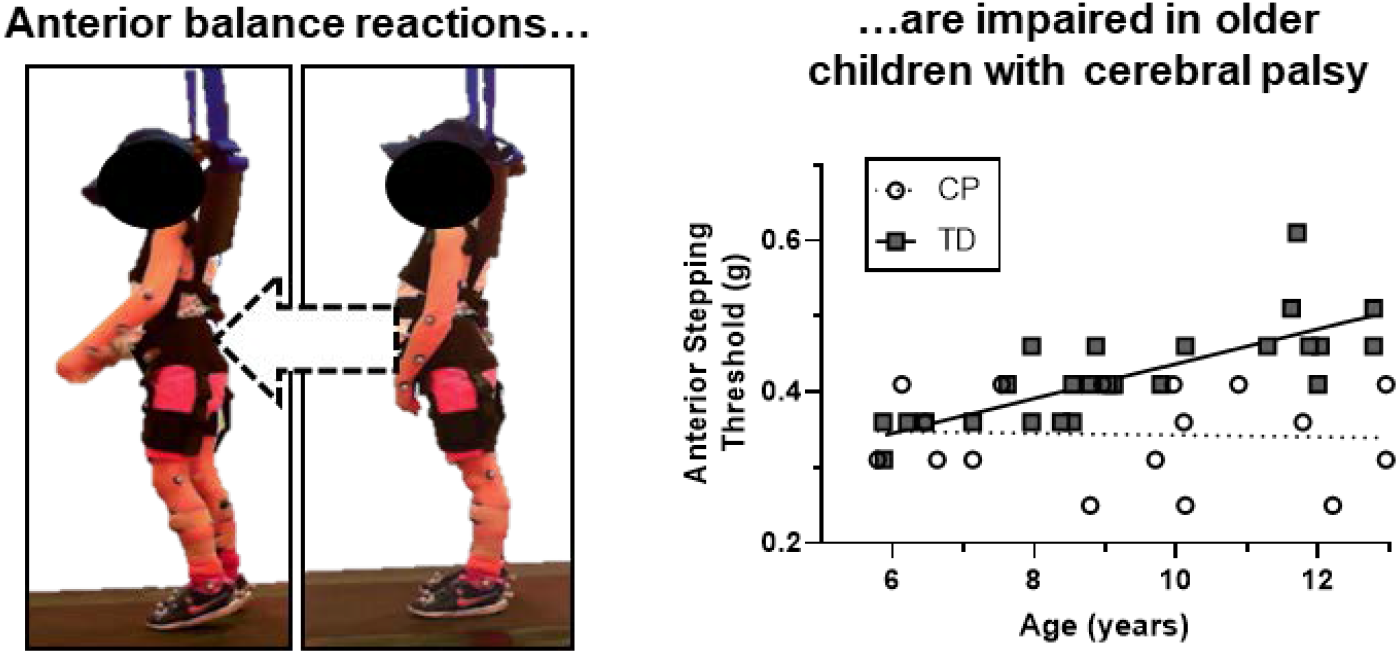

---

Falls are prevalent in ambulatory children with cerebral palsy (CP). For those with high motor function (i.e. Gross Motor Functional Classification System Levels (GMFCS) I or II), more than 75% report one or more falls per month (1). More than half of children with the highest function (GMFCS I) fall at least once per week. For those with lower function (GMFCS II), more than half fall daily. Falls lead to serious injury, accounting for 41% of femoral fractures in children with CP (2). The high risk of falls increases fear of falling, in turn limiting physical activity (3). Therefore, addressing falls in this population is key to reducing injury and enabling mobility.

The fall risk of children with CP is likely due, in part, to impaired balance reactions. Children with CP exhibit a delayed, altered coordination of the muscle and kinetic response to standing perturbations (4–7). Compared to typically developing children, those with CP may not scale their lower-extremity muscle response to the perturbation size (6), perhaps due to a limited capacity to activate muscles (6,7). The typical distal-to-proximal response to a surface translation is disrupted and more variable (4,5,7), with co-contraction being prevalent in those with CP (4,7,8). These altered neuromuscular responses likely reduce the capacity of those with CP to recover stability after a perturbation.

Limitations in balance reaction capabilities of children with CP are evident by smaller anterior single-stepping thresholds, or the perturbation magnitudes that elicit a forward step, as compared to typically developing peers (5,9). To our knowledge, *posterior* single-stepping thresholds have not been assessed in children with CP. Anatomically, anterior and posterior perturbations are characterized by different base of support dimensions, agonist torque-producing capabilities, and joint range of motions for counter-rotation movements. In older adults, another population at high risk of falls, posterior, but not anterior thresholds have been associated with future falls (10), with only moderate age-adjusted correlations between anterior and posterior thresholds (*r*=0.29) (11). Furthermore, anterior and posterior falls have distinctly different risks of impact to common fracture locations (12). Given this evidence that posterior balance reactions have distinct constraints and relevance from that of anterior responses, it is reasonable to consider how CP affects balance reactions in the posterior direction. In response to a posterior loss of balance, children with CP have an altered neural coordination between lower-extremity joints, co-contraction within the ankle joint, and a diminished kinetic output (4). So, we anticipate that posterior stepping thresholds would also be reduced in this population.

Although the neural and kinetic response to standing perturbations have been studied in children with CP (4–8), less is known about how these responses affect stability. In the context of this study, stability is determined by the horizontal trajectory of the whole-body center of mass (CoM) relative to its base of support. In a previous study that applied near-threshold perturbations inducing an anterior loss of balance, there were no differences between children with and without CP in the scaled distance between the CoM and the center of pressure (5). Given that thresholds were at smaller perturbations for those children with CP, this lack of group differences in a stability measure suggests that lower perturbation magnitudes were needed to elicit instability in children with CP. Balance reactions involve substantial velocity of the CoM relative to the base of support. So, we suggest that a measure of *dynamic* stability is appropriate in this context. The margin of stability (MoS) is a measure that accounts for the whole-body CoM position and velocity relative to the edge of the base of support (13,14). The MoS is proportional to the impulse needed to change stability states, so the measure has explicit biomechanical meaning regarding stability. To date, the MoS has not been used to evaluate perturbation responses in children with CP. Doing so can provide insight on how the resulting stability of the perturbation aligns with observed differences in balance-reaction performance.

The purpose of this study was to compare anterior and posterior single-stepping thresholds of children with and without CP. We hypothesized that children with CP would have lower thresholds in both directions. In order to gain insight on underlying mechanisms of this hypothesis, we evaluated the resulting MoS of the response, as well as the ankle plantar- and dorsiflexor activity.

## Methods

A convenience sample of 21 children with spastic CP and 29 typically developing children were enrolled in this observational, cross-sectional study. Participants were recruited regionally with advertisements, by contacting participants of previous, non-balance-related studies, and through coordination with Nemours/A.I. duPont Hospital for Children. All participants were 5-12 years of age and were able to walk without assistance from a cane or walker. We attempted to recruit in a manner so that CP and typically developing groups had representation within each combination of sex and two-year age strata. Participants were excluded if they reported any genetic, cardiovascular, metabolic, skeletal, or neuromuscular disorders (other than CP) or injuries altering mobility, balance, or safe participation. Participants were also excluded if they presented with acute illness or open lesions, had surgeries within one year to the lower-extremities, back, or shoulders, used a Baclofen pump, or were unable to follow instructions as reported by the guardian upon recruitment. Two participants with CP regularly wore bilateral ankle-foot orthoses, and were permitted to wear these devices during data collections in order to limit the risk of ankle injury. The University of Delaware institutional review board approved this study. Participants provided informed assent, and guardians provided informed consent.

This report focuses on results from balance-reaction tests, which represent part of a larger, single-session protocol more broadly assessing balance, gait, and physical activity. Previous reports that include data from some or all of these participants focused on the relationships of age, standing sway, and balance reactions in typical development (15), gait stability in children with and without CP (16), magnetic resonance elastography (MRE) to evaluate brain stiffness of the two groups (17), and the relationships between balance reaction performance and brain stiffness in children with CP (*In Review*).

All balance-reaction tests occurred in a motion analysis laboratory at the University of Delaware. Of the recruited participants, 17 children with CP (8 boys/9 girls; 13 GMFCS level I, 4 GMFCS level II; mean (SD) age=9 years, 4 months (2 years, 5 months); BMI 16.5 (1.4) kg**·**m^-2^) and 28 children with typical development (13 boys/15 girls; age=9 years, 2 months (2 years, 2 months); BMI 17.1 (2.3) kg**·**m^-2^) completed the balance-reaction test. Two children with CP and one typically developing child were unable to follow directions sufficiently for the assessment, as determined by the study team. Two participants with CP elected to end the test early due to non-physical distress—an adverse, yet anticipated outcome associated with this protocol.

The single-stepping threshold test is reported in detail in previous publications (15,18). Participants stood on a computer-controlled treadmill (ActiveStep, Simbex®, Lebanon, NH, USA, Figure 1A). They wore a safety harness (DMM, Wales, UK) attached to an overhead rail, adjusted to prevent knee or hand contact with the treadmill. Participants were instructed to “try not to step” in response to rapid, 400 ms treadmill-belt translations. After the first perturbation with an initial acceleration of 0.5 m/s^2^, a progressively challenging series of perturbations was applied. Belt accelerations inducing anterior and posterior sway were delivered in an unpredictable order. For subsequent perturbations in a given direction, the acceleration was increased by ±0.5 m/s^2^ or maintained at the previously applied magnitude, depending on success or failure. Failure was defined as either taking a step or using the harness to support more than 20% body weight (19), as recorded by a force transducer (Dillon, Fairmont, MN, USA). Anterior and posterior single-stepping thresholds were identified as the perturbation magnitude that resulted in four consecutive failed responses. In this report, “posterior” and “anterior” refer to the direction of induced sway. Thresholds were scaled to unitless values by dividing accelerations by gravity (9.81 m/s^2^) (20). Participants received a mean (SD) of 25.0 (5.1) perturbations (range=16-37).

**Figure 1.**
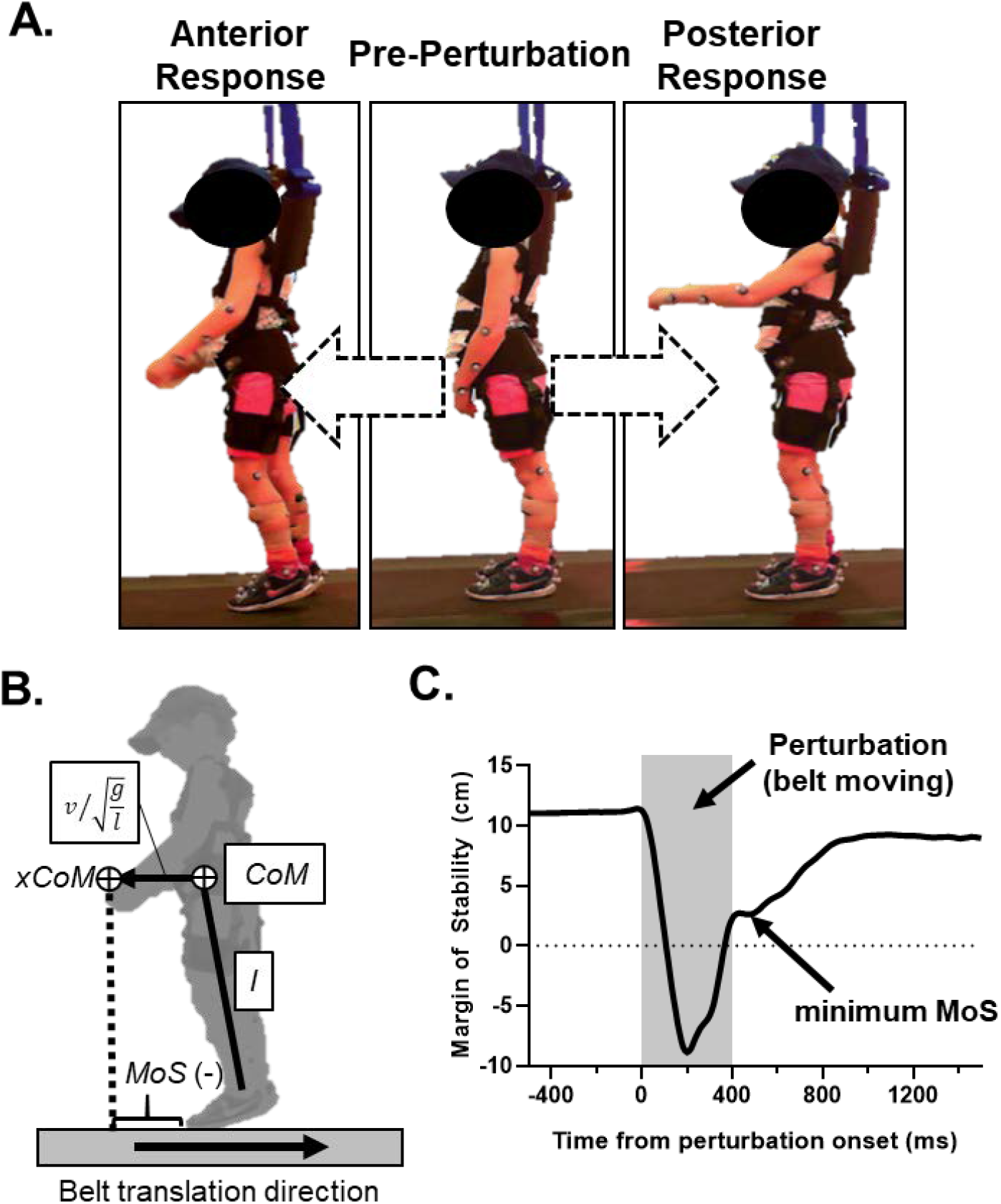
**A**. A six-year-old child with cerebral palsy responds to anterior (left, acceleration=0.20 g) and posterior (right, acceleration=0.15 g) perturbations. Consent was obtained to use these images. **B**. A model representing the margin of stability (MoS) calculation for an anterior response. The extrapolated center of mass (xCoM) location is comprised of the center of mass (CoM) position and scaled velocity 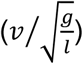. Here, velocity (*v*) is that of the CoM relative to the anterior edge of the base of support (i.e. the toe marker). Gravity (*g*) is 9.81 m/s^2^, and *l* is an estimated pendulum length based on the ankle center location and the CoM position. The MoS, then, is the distance between the vertical projection of the xCoM and the edge of the base of support. In this image, the MoS is negative, denoting an unstable state. **C**. A time series of the MoS during a successful, feet-in-place perturbation response. Analyses were conducted on the minimum MoS occurring after the end of the perturbation. We did not analyze the true minimum MoS occurring during the perturbation, as that value is directly influenced by the treadmill belt velocity, and the stabilizing phase corresponds with deceleration of the treadmill belt to a greater extent than the active stabilizing response from the child.

All participants were outfitted with 41 retro-reflective markers placed on their extremities, pelvis, trunk, and head. Marker trajectories were recorded by 12-13 cameras (Motion Analysis Corporation, Rohnert Park, CA, USA, replaced mid-study by Qualisys, Gothenburg, Sweden; 120 Hz), from which the MoS (Figure 1B) was quantified using custom LabVIEW programs (National Instruments, Austin, TX, USA) as follows:

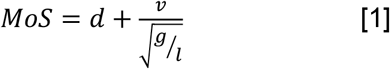

where *d=x*_*toe*_ *– x*_*CoM*_ (*x* is the anteroposterior position) and *v=v*_*toe*_ *– v*_*CoM*_ (*v* is velocity) for anterior perturbations, and *d=x*_*CoM*_ *– x*_*heel*_ and *v=v*_*CoM*_ *-v*_*heel*_ for posterior disturbances. The CoM position was calculated from kinematic data and anthropometric values (21). The variable *g* represents gravity (9.81 m/s^2^) and *l* is the sagittal-plane distance between the mean ankle joint center location and that of the whole-body CoM, calculated on a frame-by-frame basis. For feet-in-place responses analyzed in this study, we identified the minimum MoS from the point after which the belt translation ended (Figure 1C). We chose the more positive (i.e. more stable) minimum MoS between the left and right limbs. Then, to scale the MoS to a unitless value, it was divided by body height. In order to understand how stability was maintained after a challenging perturbation, we evaluated the minimum MoS of the largest perturbation after which a feet-in-place response was maintained. In order to understand how stability was maintained after a common perturbation magnitude, we assessed the response to anterior and posterior perturbations of 0.20 g (i.e. the 4^th^ smallest perturbation) and 0.10 g (i.e. the 2^nd^ smallest perturbation), respectively. These common levels allowed us to include all but one typically developing participant in the assessment of anterior perturbations, and all but five participants (2 CP, 3 typically developing) in the assessment of posterior perturbations.

Muscle activity was recorded using surface electromyography (EMG; Delsys, Natick, MA, USA; 1200 Hz) on the bilateral medial gastrocnemius and tibialis anterior (22). The EMG signals were filtered with a 1^st^-order polynomial, band-pass filter (10-300 Hz). Signals were then de-meaned, rectified, and low-pass filtered (8th order Butterworth) at 50 Hz for muscle onset latency and 4 Hz for peak activation and co-contraction ratios. A higher frequency cutoff was used for muscle onset latency due to the effect of filtering on time-based measures of muscle activity (23). All muscle activity was scaled to the median activity of a 500 ms period before perturbation onset. In our analyses of muscle activity, the agonists refer to the muscle resisting the sway induced by the perturbation. So, for anterior perturbations (i.e. posterior translations that induce anterior sway), the agonist is the gastrocnemius and the antagonist is the tibialis anterior. Muscle onset latency was defined as the time after perturbation onset at which scaled muscle activity of the agonist exceeded three standard deviations above the median pre-perturbation activity, with that activity sustained for at least 200 ms. Peak muscle activation was the maximum agonist amplitude achieved during the perturbation response. The co-contraction ratio (agonist/antagonist) was that of the integral of each muscle’s activity within a 500 ms period after perturbation onset. Outliers in these variables, defined as those values more than three times the interquartile range from the median, were removed before statistical analyses. As with the MoS analysis, we assessed EMG results at near-threshold and common perturbation levels. The first three children with CP, as well as the first six children with typical development, were part of an early pilot and feasibility study of the protocol reported here. These nine participants were not instrumented with EMG, as that measure was added later. As a result of a smaller sample outfitted with EMG (12 CP, 22 typically developing), fewer samples for the common-level analysis, and our removal of outliers (CP: 0-3 outliers; TD: 0-5 outliers), EMG analyses were conducted on 9-12 CP participants and 17-22 typically developing participants, depending on the dependent variable.

Recruitment of 25 children with CP and 25 typically developing children was planned to achieve 80% power to detect a large between-group difference (Glass’s Δ=0.80, α=0.05). However, early challenges in recruiting CP participants limited our ability to meet this goal within our timeline. Furthermore, in reviewing our data, we noted that age likely played a meaningful role as a covariate. So, we used general linear models to assess the effects and interactions of group (CP or typically developing) and age on single-stepping thresholds and the minimum MoS. In order to assess EMG variables, we also included the within-subjects factor of limb in our full factorial model. Right and left limbs were identified as dominant (i.e. self-reported kicking limb) or non-dominant. In the case of significant interactions including age, post-hoc analyses with Sidak adjustments were conducted at estimated marginal means of relatively younger and older ages of 7 and 11 years, respectively. In order to reduce the number of statistical tests, we only evaluated the MoS and EMG variables in the case of between-group differences in anterior or posterior thresholds. Statistics were evaluated using SPSS (v24, IBM, Armonk, NY, USA). With our achieved sample size of 45 participants, we had 75% power to detect a large main effect or interaction (partial η^2^ ≥ 0.14; α<0.05) of age and group in our stepping thresholds.

## Results

A significant interaction of age and group (p=0.001, partial η^2^=0.24) revealed that anterior thresholds increased with age in typical development to a greater extent than in children with CP (Figure 2A). At 7-year age estimates, typically developing children did not have higher thresholds than their CP peers (p=0.33, partial η^2^=0.02). However, at 11 years, typically developing children demonstrated larger thresholds (p<0.001, partial η^2^=0.48).

**Figure 2.**
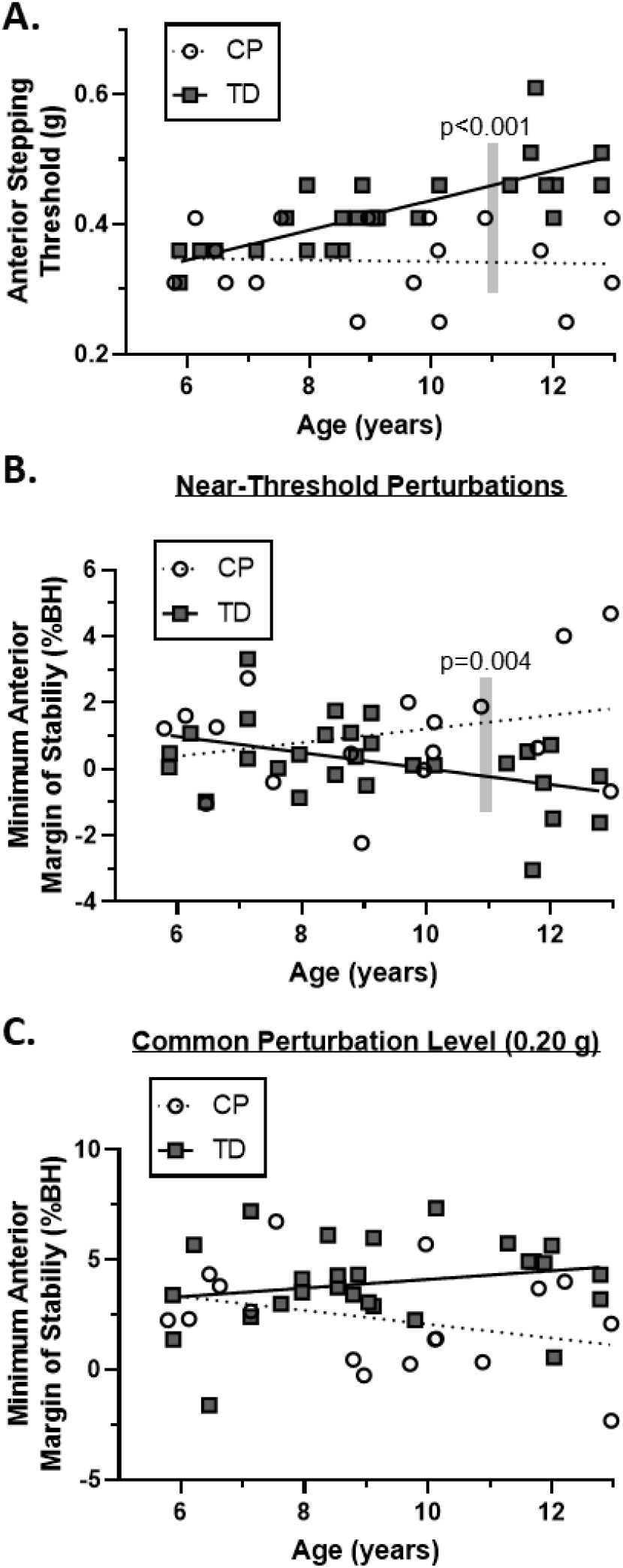
**A**. Anterior single-stepping thresholds (in units of gravity) of children with cerebral palsy (CP, open circles and dashed line) and typically developing children (TD, solid squares and line) as a function of age. A significant between-group difference at an estimated age of 11 years is denoted with a gray bar. **B**. The minimum margin of stability, as a percentage of body height, after near-threshold-level perturbations (i.e. one level below the threshold level that induced a step). A significant between-group difference at an estimated age of 11 years is denoted with a gray bar. **C**. The minimum margin of stability at a common perturbation size of 0.20 g.

Dynamic stability analyses of anterior balance reactions confirmed that between-group differences in stability control were most profound at older ages. At near-threshold levels, a significant interaction of age and group (p=0.02, partial η^2^=0.12) on the minimum MoS was apparent (Figure 2B). At 7-year age estimates, between-group differences were small (p=0.80, partial η^2^=0.002). However, at 11 years of age, typically developing children demonstrated dynamic instability, while children with CP had positive minimum stability values (p=0.004, partial η^2^=0.18). At a common perturbation level of 0.20 g, the interaction of age and group on the minimum MoS had a medium effect size (p=0.08, partial η^2^=0.07). Here the trend was such that older children with CP were less stable after the perturbation compared to older, typically developing children (Figure 2C). At this perturbation size, no main effects of age (p=0.68, partial η^2^=0.004) or group (p=0.27, partial η^2^=0.03) were large.

There were few group differences in the magnitude, timing, or co-contraction associated with the ankle muscle response to an anterior perturbation. At near-threshold levels, a significant interaction of limb, group, and age (p=0.03, partial η^2^=0.17) was apparent for the maximum medial gastrocnemius activity. In post-hoc analyses, the only notable between-group difference was observed for the dominant limb at an estimated age of 7 years, with typically developing children having more activity of the medial gastrocnemius (p=0.01, partial η^2^=0.23). At near-threshold levels, no main effects or interactions containing group were large (p=0.28-0.81, partial η^2^=0.002-0.04) for the onset latency or co-contraction index. At a common perturbation level, no large main effects or interactions containing group were observed for any EMG variable (p=0.23-0.93, partial η^2^<0.054). All EMG data are presented as supplementary material.

Posterior stepping thresholds were larger with age (p=0.01, partial η^2^=0.17; Figure 3). Main effects (p=0.37, partial η^2^=0.02) and interactions (p=0.17, partial η^2^=0.05) including group were not significant. The medium-sized effect of the group and age interaction suggests that we may not have detected a trend in which between-group differences in posterior balance reactions were more apparent for the older children. As per our planned statistical approach, we did not analyze MoS or EMG variables. These data, however, are presented as supplementary material.

**Figure 3.**
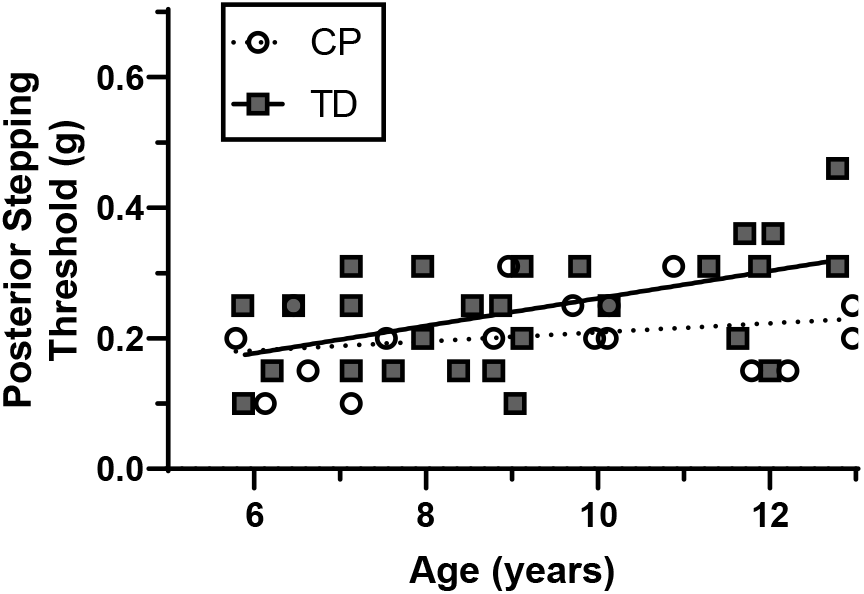
Posterior single-stepping thresholds (in units of gravity) of children with cerebral palsy (CP, open circles and dashed line) and typically developing children (TD, solid squares and line), as a function of age.

## Discussion

The purpose of this study was to compare anterior and posterior single-stepping thresholds of children with and without CP. We hypothesized that children with CP would have lower thresholds in both directions. This hypothesis was partially supported, as we observed large between-group differences in anterior, but not posterior, thresholds. Between-group differences were most evident in the older children.

Previous studies have also demonstrated between-group differences in anterior stepping thresholds (5,9), yet smaller samples prevented consideration of an age and group interaction. A study by Chen and colleagues (5) demonstrated that children with CP (age 8-13 years) had lower anterior thresholds than age-matched typically developing children *and* younger (4-7 years) typically developing children. This result can be interpreted as the negative effects of both pathology and developmental delays. In a more rigorous study design that included younger typically developing children, our results suggest that anterior balance-reaction impairments were primarily influenced by developmental delays. This interpretation is based on our observation of no between-group differences in stepping thresholds at our lower age range. This lack of group differences does not align with that of Burtner and colleagues (9), a discrepancy likely due to the high-functioning status of our CP group (GMFCS levels I and II) compared to that reported previously (levels I-III). We anticipate that lower-functioning children with CP would have lower thresholds at a young age. However, we do not know if the trajectory of this skill with age would remain flat or decline. Generally, the gross-motor function of children at GMFCS Levels I and II plateaus in early childhood (24,25). But, for GMFCS Levels III and IV, that function can decline with age after reaching an early peak value. So, we hypothesize that balance reaction capabilities in lower-functioning participants may also decline with age.

Dynamic stability analyses revealed that, in response to the most challenging of anterior perturbations, the older, typically developing children recovered from instability while their CP counterparts did not (Figure 2B). We cannot determine if this result is due to group differences in the ability to prevent a step when unstable, or a preference of CP participants to conservatively step despite a stable state. The MoS measure is calculated under the false assumption that the center of pressure, as modulated by the ankle flexors and extensors, can instantaneously be positioned at the edge of the base of support (26). So, any delay in generating a plantarflexor moment and translating the center of pressure underneath the base of support would limit the minimum MoS value that could be maintained without a step. Previous studies have suggested that children with CP have a delayed and slower ankle torque generation after a perturbation (5), as well as an altered, slower trajectory of the center of pressure (9). Given this information, we suggest that children with CP have altered MoS limits, more so than a conservative preference for stepping when not necessary. At common anterior perturbation magnitudes, the interaction of age and group on the minimum MoS was not as strong, with a trend suggesting that the perturbation was more destabilizing in older children with CP (Figure 2C). This lack of a strong group effect underscores the need to evaluate balance reactions at a challenging level for each participant (9). We suggest that the common level of 0.2 g was too benign to necessitate a maximum stabilizing effort in the more-skilled participants.

The latency, magnitude, and co-contraction of the ankle musculature did not show trends that aligned with the group differences in anterior stepping thresholds. This result does not agree with previous evidence of a delayed, altered coordination of the muscle response in children with CP (4,6,7). Key limitations to our study include a limited sample size for our EMG analyses; however, we did not detect effect sizes worth pursuing in a larger study. Additionally, our high-functioning group of children with CP may not have the altered neural responses of the more impaired participants in previous studies. Furthermore, we did not measure muscle responses about the knee and hip, two areas where distinct responses in children with CP have been observed (8), including an altered coordination between distal and proximal joints (4,5,7). Lastly, our participants with CP did not exhibit notable crouch while standing, an aspect that may directly underlie previous between-group differences in the neural response to a perturbation (8).

The effects of CP and age on posterior balance reactions were less profound than that in the anterior direction. Perhaps group differences in posterior thresholds would become more evident at older ages. A positive trajectory of posterior thresholds with age in typical development (Figure 3) (15) suggests that this skill is developing. It is reasonable to suggest that limited plantarflexor strength associated with CP (27) is a factor underlying group differences in anterior thresholds (6,7). Given that the tibialis anterior generates much smaller ankle moments than that of the plantarflexors (27), it may be that posterior thresholds are influenced more by reaction time than torque-generating capacities. Of note, there did not appear to be between-group differences in tibialis anterior latencies in response to posterior perturbations (see supplementary material). General reaction time improves with typical development up to the age of 16-17 years (28). If reaction time is a major underlying factor of posterior feet-in-place balance reactions, then we may expect posterior thresholds to continue to improve with age in typical development, eventually leading to between-group disparities in performance.

Our findings suggest that high-functioning CP is associated with an age-related and direction-specific impairment in balance reaction capabilities. These impairments aligned with differences in stability maintenance after the perturbation, but were not associated with ankle muscle activation characteristics. We do not know if our observations are specific to the means of applying the perturbation (e.g. surface translation, waist pull, or lean-release), nor do we know how CP may affect balance reactions in other context (e.g. lateral perturbations or perturbations during gait). Our results can guide hypotheses for more thorough, longitudinal studies on how development and baseline function alter the trajectory of balance reaction skill. Of note, balance reactions can be improved with practice in children with CP (7). Thus, the capabilities that we’ve quantified here can serve as a modifiable target for interventions to improve that skill and, potentially, alter the risk of falling.

## Data Availability

Data is not available for sharing.

## Abbreviations

GMFCS: Gross Motor Functional Classification System Levels
CoM: center of mass
MoS: margin of stability
CP: cerebral palsy
TD: typically developing

## Acknowledgements and Funding

This work was supported by the Delaware INBRE program with a grant from the NIGMS (P20-GM103446) and the State of Delaware. Also supported by the Eunice Kennedy Shriver NICHD 1R01HD090126. Subject recruitment and scheduling were made possible with resources provided by the Delaware Rehabilitation Institute.

## Supplementary Materials

**Figure S1.**
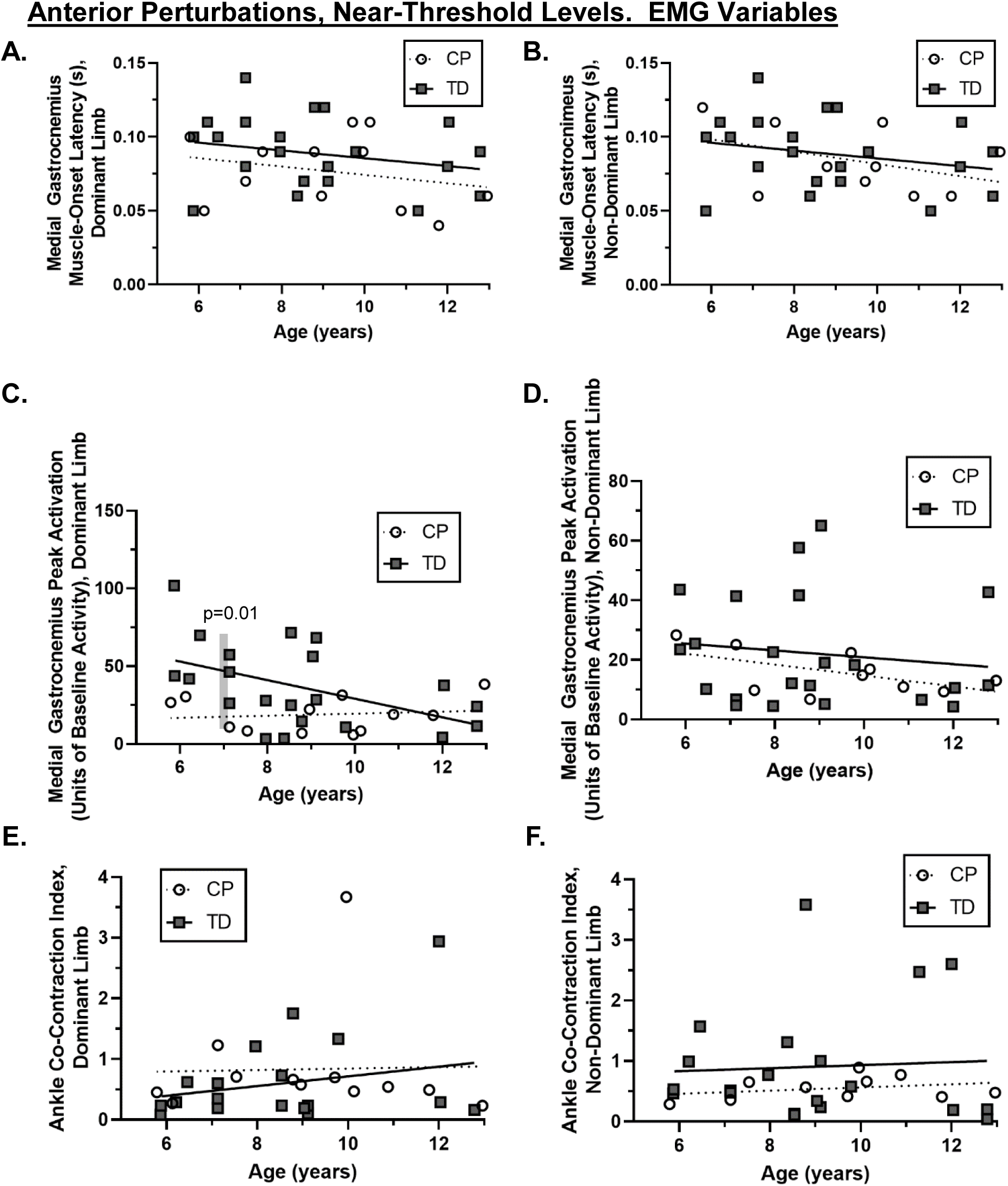
EMG variables for the response to anterior perturbations at near-threshold perturbation magnitudes. EMG variables include the muscle onset latency (A,B), peak activation (C,D), and co-contraction index (E,F) of the dominant (A,C,E) and non-dominant (B,D,F) limbs. A significant between-group difference in dominant-limb peak activation at an estimated age of 7 years is noted with a shaded region (C).

**Figure S2.**
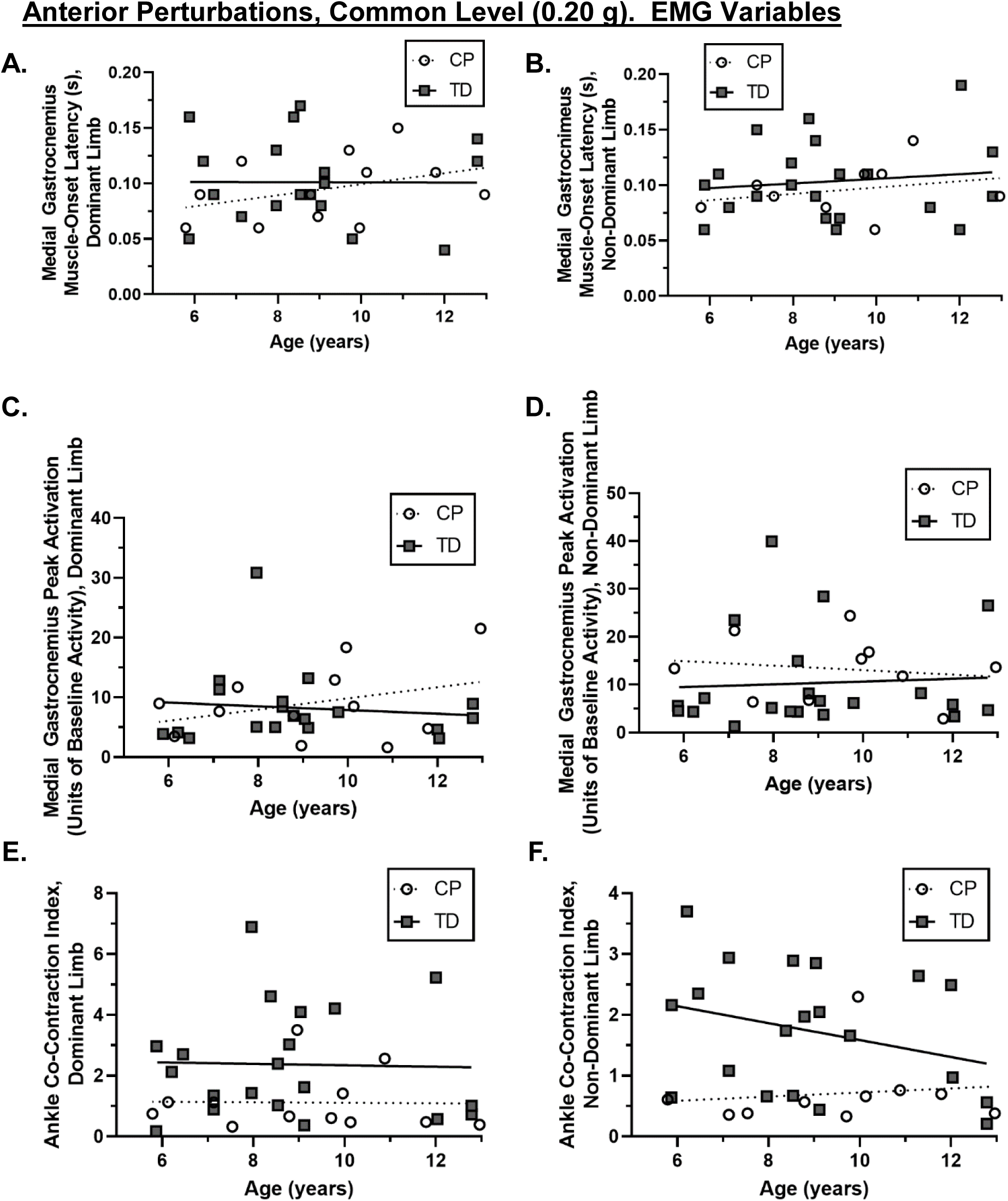
EMG variables for the response to anterior perturbations at a common perturbation magnitude of 0.20 g. EMG variables include the muscle onset latency (A,B), peak activation (C,D), and co-contraction index (E,F) of the dominant (A,C,E) and non-dominant (B,D,F) limbs.

**Figure S3.**
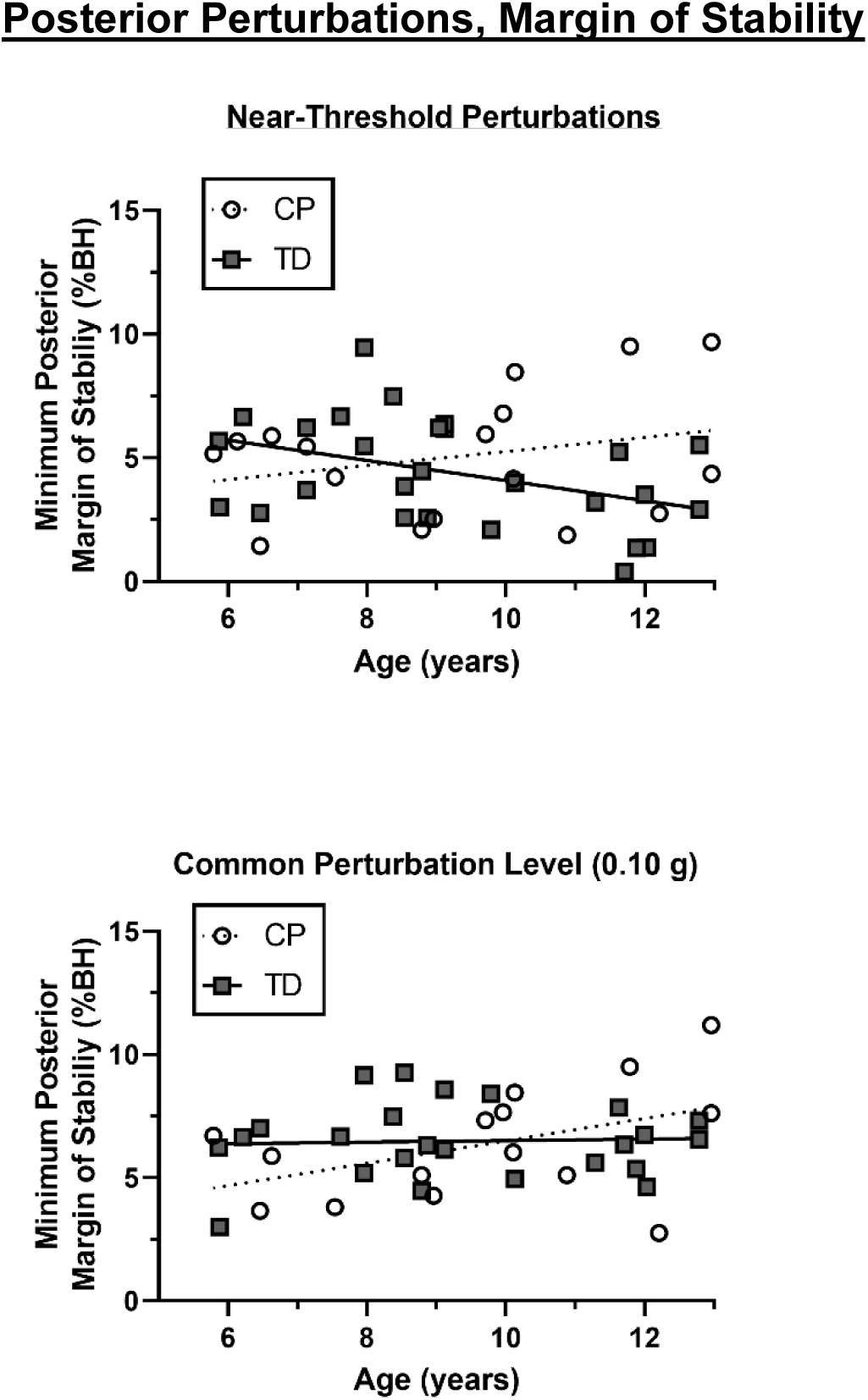
Minimum margin of stability variables for the response to posterior at near-threshold perturbation magnitudes (A) and at a common perturbation magnitude of 0.10 g (B). As per our analysis plan, we did not conduct statistical analyses on these variables, as no between-group differences in posterior stepping thresholds were observed.

**Figure S4.**
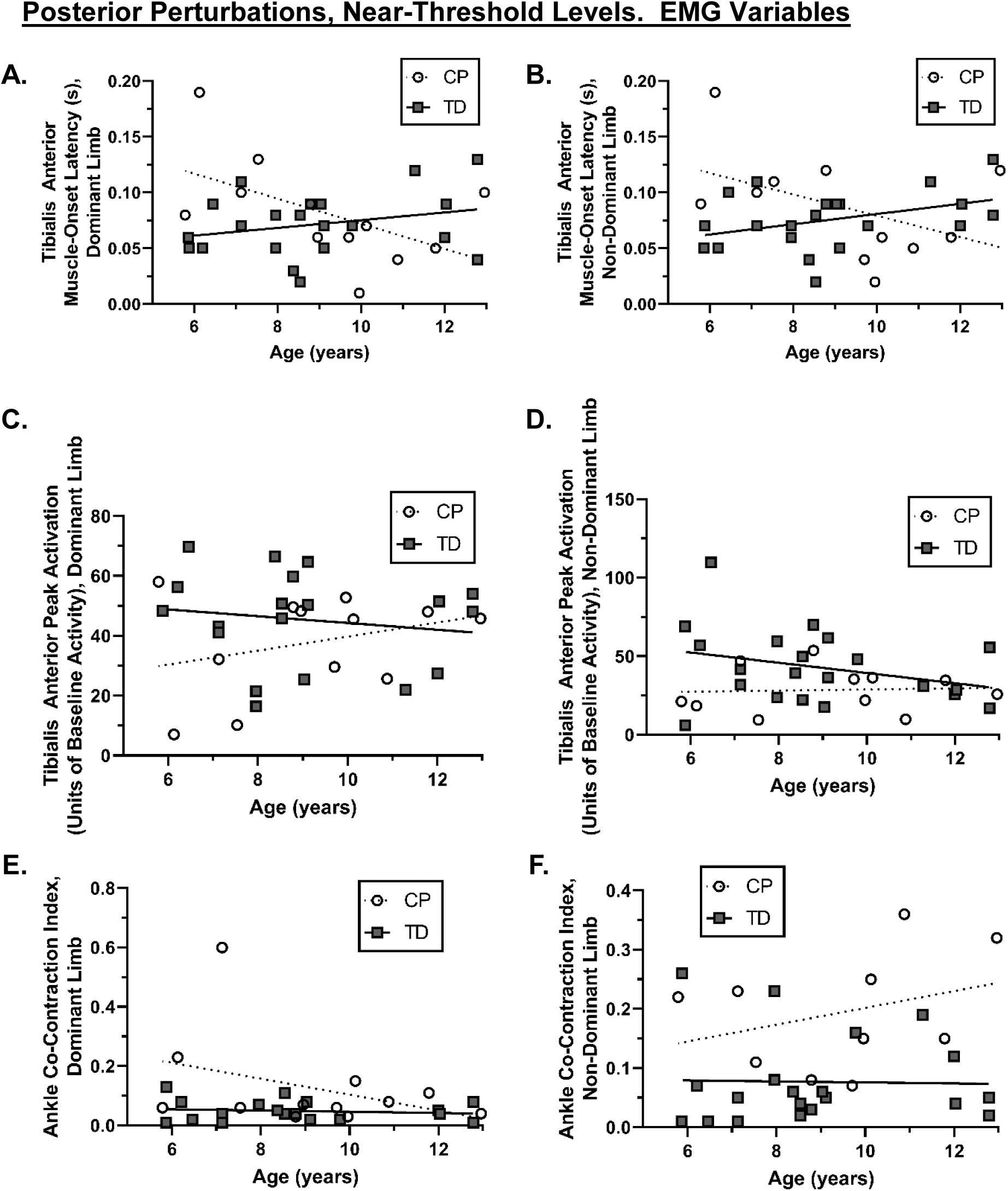
EMG variables for the response to posterior perturbations at near-threshold perturbation magnitudes. EMG variables include the muscle onset latency (A,B), peak activation (C,D), and co-contraction index (E,F) of the dominant (A,C,E) and non-dominant (B,D,F) limbs. As per our analysis plan, we did not conduct statistical analyses on these variables, as no between-group differences in posterior stepping thresholds were observed.

**Figure S5.**
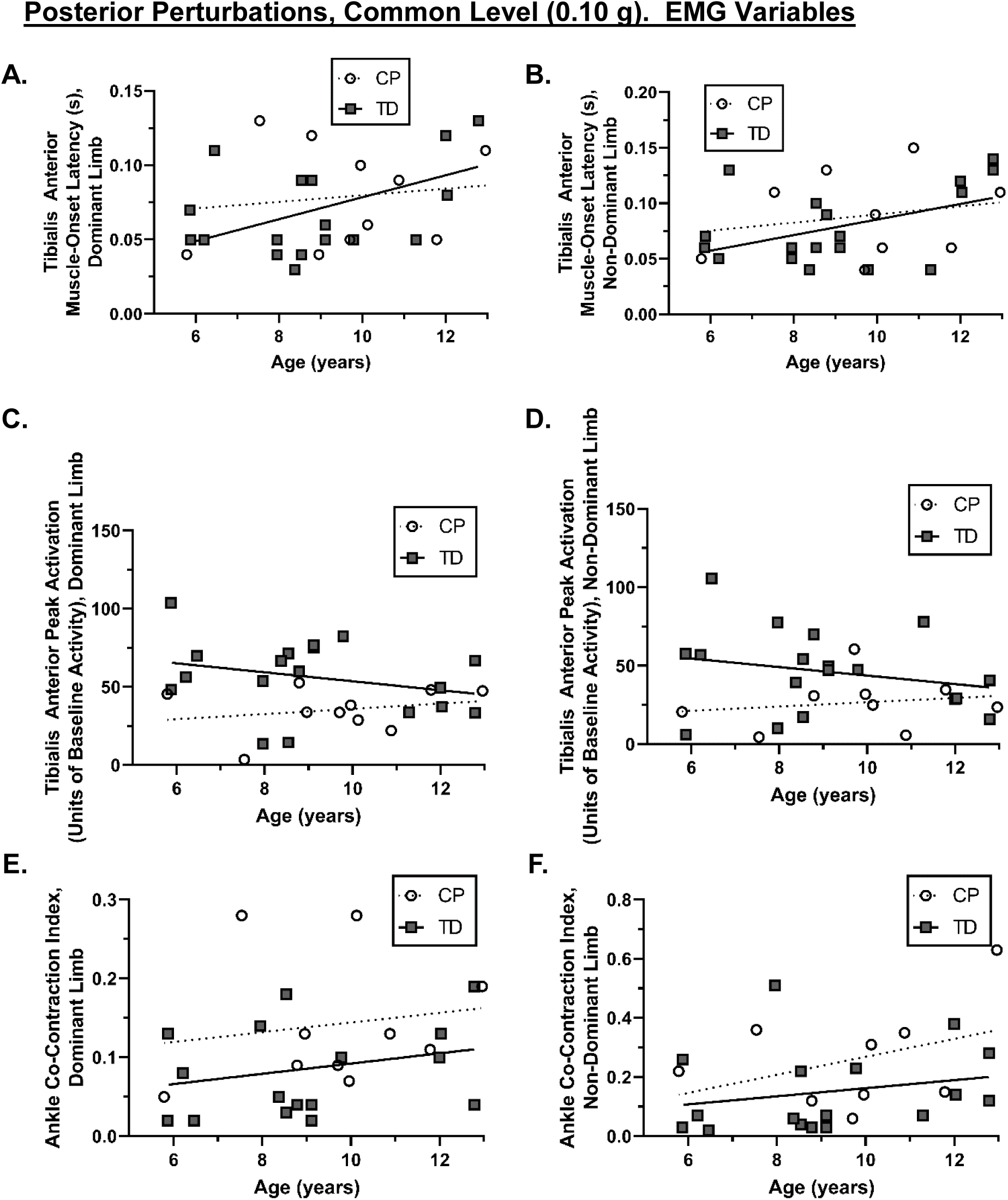
EMG variables for the response to posterior perturbations at a common perturbation magnitude of 0.10 g. EMG variables include the muscle onset latency (A,B), peak activation (C,D), and co-contraction index (E,F) of the dominant (A,C,E) and non-dominant (B,D,F) limbs. As per our analysis plan, we did not conduct statistical analyses on these variables, as no between-group differences in posterior stepping thresholds were observed.

## References

1. Boyer ER, Patterson A. Gait pathology subtypes are not associated with self-reported fall frequency in children with cerebral palsy. Gait Posture. 2018;63:189–94.

2. Loder RT, O’Donnell PW, Feinberg JR. Epidemiology and mechanisms of femur fractures in children. J Pediatr Orthop. 2006;26:561–6.

3. Verschuren O, Wiart L, Hermans D, Ketelaar M. Identification of facilitators and barriers to physical activity in children and adolescents with cerebral palsy. J Pediatr. 2012;161:488–94.

4. Nashner LM, Shumway-Cook A, Marin O. Stance posture control in select groups of children with cerebral palsy: deficits in sensory organization and muscular coordination. Exp Brain Res. 1983;49:393–409.

5. Chen J, Woollacott MH. Lower extremity kinetics for balance control in children with cerebral palsy. J Mot Behav. 2007;39:306–16.

6. Roncesvalles MN, Woollacott MW, Burtner P. Neural factors underlying reduced postural adaptability in children with cerebral palsy. Neuroreport. 2002;13:2407–10.

7. Woollacott MH, Shumway-Cook A. Postural dysfunction during standing and walking in children with cerebral palsy: What are the underlying problems and what new therapies might improve balance? Neural Plast. 2005;12:211–9.

8. Burtner P, Qualls C, Woollacott M. Muscle activation characteristics of stance balance control in children with spastic cerebral palsy. Gait Posture. 1998;8:163–74.

9. Burtner PA, Woollacott MH, Craft GL, Roncesvalles MN. The capacity to adapt to changing balance threats: a comparison of children with cerebral palsy and typically developing children. Dev Neurorehabil. 2007;10:249–60.

10. Sturnieks DL, Menant J, Delbaere K, Vanrenterghem J, Rogers MW, Fitzpatrick RC, et al. Force-controlled balance perturbations associated with falls in older people: A prospective cohort study. PLoS One. 2013;8:e70981.

11. Crenshaw JR, Bernhardt KA, Atkinson EJ, Khosla S, Kaufman KR, Amin S. The relationships between compensatory stepping thresholds and measures of gait, standing postural control, strength, and balance confidence in older women. Gait Posture. 2018;65:74–80.

12. Crenshaw JR, Bernhardt KA, Achenbach SJ, Atkinson EJ, Khosla S, Kaufman KR, et al. The Circumstances, Orientations, and Impact Locations of Falls in Community-Dwelling Older Women. Arch Gerontol Geriatr. 2017;73:240–7.

13. Hof AL, Gazendam MGJ, Sinke WE. The condition for dynamic stability. J Biomech. 2005;38:1–8.

14. Crenshaw JR, Grabiner MD. The influence of age on the thresholds of compensatory stepping and dynamic stability maintenance. Gait Posture. 2014;40:363–8.

15. Conner BC, Petersen DA, Pigman J, Tracy JB, Johnson CL, Manal K, et al. The cross-sectional relationships between age, standing static balance, and standing dynamic balance reactions in typically developing children. Gait Posture. 2019;73:20–5.

16. Tracy JB, Petersen DA, Pigman J, Conner BC, Wright HG, Modlesky CM, et al. Dynamic stability during walking in children with and without cerebral palsy. Gait Posture. 2019;72:182–7.

17. Chaze CA, McIlvain G, Smith DR, Villermaux GM, Delgorio PL, Wright HG, et al. Altered brain tissue viscoelasticity in pediatric cerebral palsy measured by magnetic resonance elastography. NeuroImage Clin. 2019;

18. Crenshaw JR, Kaufman KR. The intra-rater reliability and agreement of compensatory stepping thresholds of healthy subjects. Gait Posture. 2014;39:810–5.

19. Cyr M-A, Smeesters C. Maximum allowable force on a safety harness cable to discriminate a successful from a failed balance recovery. J Biomech. 2009;42:1566–9.

20. Hof AL. Scaling and Normalization. In: Wolf SI, Muller B, editors. Handbook of Human Motion. New York: Springer; 2018. p. 295–305.

21. Winter DA. Anthropometry. In: Biomechanics and Motor Control of Human Movement. Fourth. New Jersey: John Wiley & Sons, Inc.; 2009. p. 82–106.

22. Hermens HJ, Freriks B, Disselhorst-Klug C, Rau G. Development of recommendations for SEMG sensors and sensor placement procedures. J Electromyogr Kinesiol. 2000;10:361–74.

23. Di Fabio RP. Reliability of computerized surface electromyography for determining the onset of muscle activity. Phys Ther. 1987;67:43–8.

24. Smits DW, Gorter JW, Hanna SE, Dallmeijer AJ, Van Eck M, Roebroeck ME, et al. Longitudinal development of gross motor function among Dutch children and young adults with cerebral palsy: An investigation of motor growth curves. Dev Med Child Neurol. 2013;55:378–84.

25. Hanna SE, Rosenbaum PL, Bartlett DJ, Palisano RJ, Walter SD, Avery L, et al. Stability and decline in gross motor function among children and youth with cerebral palsy aged 2 to 21 years. Dev Med Child Neurol. 2009;51:295–302.

26. Hof AL, Curtze C. A stricter condition for standing balance after unexpected perturbations. J Biomech. 2016;49:580–5.

27. Engsberg JR, Ross SA, Olree KS, Park TS. Ankle spasticity and strength in children with spastic diplegic cerebral palsy. Dev Med Child Neurol. 2000;42:42–7.

28. Fuchigami T, Okubo O, Fujita Y, Okuni M, Okuni M, Noguchi Y, et al. Auditory event-related potentials and reaction time in children: evaluation of cognitive development. Dev Med Child Neurol. 1993;35:230–7.

